# What are we learning with Yoga: a text mining approach to literature

**DOI:** 10.1101/2022.12.05.22282979

**Authors:** Rosangela Ieger Raittz, Camilla R. De Pierri, Camila P. Perico, Diogo J. Machado, Jeroniza N. Marchaukoski, Roberto T. Raittz

**Author notes:** Corresponding author: Roberto T. Raittz.

## Abstract

Techniques used in yoga existed long before science as we know it. However, with Yoga westernization, the proliferation of scientific studies addressing the subject is exponential. Even though the literature presents thousands of related articles, many studies are considered flimsy — the diversity of methods, areas of interest, and focus cause it to become a complex environment without general guidelines for the researcher. This status may represent one of the impediments to the field’s evolution. This study aims to elaborate a global structure of the literature on Yoga to open the door for multidisciplinary collaboration and bring new light to some relevant aspects. Due to its complexity, we understand that techniques to model complex domains are necessary. Contemporary methods of Artificial Intelligence have promoted advances in Bioinformatics, including text mining to scientific literature. Through the vector representation of texts, we got semantic abilities and could organize data in a way that allowed us to acquire a global vision of this literature. Additionally, we made the platforms available to Yoga researchers to enable them to evaluate our findings and make their forays into the literature. Besides better understanding why we study Yoga, we discovered that the literature offers insights into the relationships in broader aspects.

## 1. Introduction

Yoga has been taught for thousands of years [1][2][3], and there is inevitably a vast cultural legacy related to it. However, uniting traditional knowledge with scientific principles and language is difficult. Experts who have the domain of Yoga knowledge don’t belong (necessarily) to the academic environment. The scientific literature on Yoga or Yoga practice (YP) contains thousands of articles and hundreds of reviews. The reported benefits related to YP have motivated the verification of new application possibilities, and the trend is that the proliferation of interest in the subject will continue to expand.

Groups develop the studies from several areas, often far apart (from biochemistry to social sciences, for instances), and most focus on their specific interests. Unfortunately, there is no consensus about a general guideline to guide multidisciplinary approaches, although there are initiatives in this direction. Thus, the accumulation of scientific knowledge about Yoga is harmed once research results are not always shared efficiently due to the difficulty of relating them. Furthermore, confidence in the results decreases because similar results are not easily comparable due to the heterogeneity of the experimental components and the lack of a one-size-fits-all approach to Yoga. Furthermore, most literature targets medical or therapeutic aspects where methodological rigor is in high demand, which may hide the value of results in other domains. On the other hand, to conciliate the various studies on different Yoga styles, it has been suggested that the term YBP (Yoga Based Practice) can be adopted in generic research [4]. Studies in [5][6] present comprehensive discussions on approaches in the Yoga area.

We found studies focusing on the construction, discussion, and recomposition of frameworks from the perspective of integration of the area [7][8]. These models often seek to relate traditional approaches to scientific arguments [9]. Other strategies target specific technical aspects of a particular area of knowledge that hinder multidisciplinary access [10][11]. Anyway, these theoretical studies are a rich source of information when the intention is to understand Yoga as a research area, and are necessary references in this sense.

Emerging technologies in information, Artificial Intelligence, and data science can elaborate and represent information in ways that capture hidden meanings and make complex relationships between concepts. However, we found only one study that approaches data science to Yoga literature. It searches for trends in future research in Yoga [2] and the emergence of studies related to the nervous system. However, it doesn’t consider semantic relations among the articles in the literature.

Vector representation approaches to represent textual documents have shown the most promise among text mining (TM) techniques [12][13][14]. Text-associated vectors are related as points in a multi-dimensional space, enabling direct comparison between two documents using geometric measures of distances [15].

This study presents an environment for text mining in Yoga literature. The TM technology allowed us to establish a synthetic view of the literature and to define consistent structural elements for the relation between the several aspects contemplated in the articles.

Our analyses showed that the articulation of the literature is helpful to understand, in a broad view, the benefits that can derive from YP and the profiles of its practitioners. However, we also found that elements could be better addressed in the studies and support a better integration of the literature. We also verified the possible evolution of theoretical frameworks aimed at instrumentalizing yoga researchers by considering the learning measure as one of the results of YP.

The information/knowledge organization applied in the elaboration applied in this article is available for public use at [16].

## 2. Methods

We focused our study on YOGA papers in the PubMed database. Aiming at a general view, the basis PubMed search was *YOGA [title/abstract] AND (“1970/1/1”[Date - Publication]* : *“2022/6/3”[Date - Publication])*, which returned 6905 papers in a table with the fields pmid, Title, Abstract, Authors, and Data. We excluded Papers in which the fields Title or Abstract were empty and those whose number of characters within these fields was less than 300. Repeated titles also were excluded. Two sets of papers were considered independently for Text Mining (TM). The first is with 5782 pieces (complete), and another considers only documents within the term YOGA in the Title, with 2929 articles (TextDoc). A text document is a set of characters composed of a title and an abstract concatenated. We will refer to the corpus in the complete TM environment as TextDocAll and that of the specific analysis TexDoc. In addition, pieces of information from other fields were conserved related to each document.

### 2.1. Documents and words vectors

The strategy we used to create the vector systems for semantic analysis consists of transforming TextDocs into a biological information format to exploit Bioinformatics tools. To construct base vectors (Wbase), we coded every document into amino acid symbols [17] and represented them in vectors using the SWeeP approach [18]. We set the projection length for SWeeP vectors as 1369, as used to vectorize whole genome sequences [19].

The word embedding adopted defines vectors representing a specific word (Wwrd) as a mean of the Wbase of TextDoc containing that word. The TextDoc embedding vectors (Wtxt) representing texts in the corpus are the mean of Wwrd for the words within each document. So:

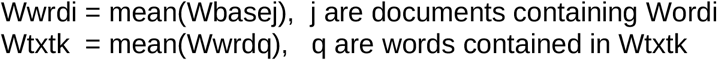

In the proposed model, words and documents are dots in the same vector space where any Word is comparable to another word or document (and vice-versa) through vector distance metrics.

We chose a subset of words (WRD) to compose the relevant words list considering the relative frequency of the words in the YOGA PubMed search to the Kaggle frequency in [20]. More specific details, such as Filtering relevant terms and disambiguating, will be omitted to avoid unnecessary complexity. The vectors (Wbase, Wwrd, and Wtxt) and correspondent lists of words (WRD) and TextDoc (TEXT) are available in supplementary. However, the complete set of words (WRD0) and respective vectors (Wwrd0) are not available, but those were used in analyses, including searching the database to build HTML-TM tools.

### 2.2. Semantic search using vectors

The presented structure permits determining which words are closest to a given query (other words or papers) in TEXT or WRD databases through vector distance comparison. In this paper, an uppercase word, or a list of words (or text codes) separated by a hyphen, represents a query for a semantic search whose correspondent vector is calculated by the average of vectors of the list to be searched. For instance, NEUROLOGICAL-BRAIN-CORTEX is a query WQ such as:

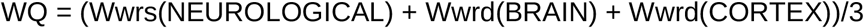

To be searched in Wwrd0, Wwrd, or Wtxt.

For some tasks, when more generalization was needed, we used a smaller set of PCA (principal component analysis) components (50-100-300) instead of the whole vector.

### 2.3. String searches and logic expressions (LOGEXP)

We define logic expressions (LOGEXP) for this study to help understand the examples involved with string searches. Operators utilized are: ‘(‘, ‘)’, ‘|’ (or), ‘&’ (and), ∼ (not), ‘_’(a space is necessary). The parameters are substrings that may occur in a TextDoc or Document Title. In some dendrograms (trees) in complementary material, the parenthesis may appear replaced by braces due to the restriction of the visualization employed tool. For instance, a vector representing the mean of documents containing the substrings ‘CARDI’ and ‘VASCUL’ is represented by (_CARDI & VASCUL). Observe in the example that the substring ‘CARDI’ must be preceded by a space to validate this match.

### 2.4. HTML-TM platforms for YOGA

For every word in the set of top related to the search (n= 2878), we searched and saved the nearest 30 correlated terms from the complete list of words (n= 20052) and the list of documents (n= 2929); we also calculated an index of the year usage for each word. An HTML-TM file containing the short seven best hits list (words), a tree of 30 top related terms, and a list of 30 closest TextDoc (papers) (WORDS.html). A second HTML-TM file (TEXTS.html) permits the user to search for similar articles. In both files, the paper numbers are consistent, facilitating navigating the literature contents quickly. These files are supplementary and accessible by loading the HTML-TM files in a browser.

### 2.5. Clusters and distances

We constructed all Hierarchic clusters with the Neighbor-Joining method for phylogenetic trees. Dendoscope [21] and ITOL [22] are the tools used for tree visualization and presentation. The Euclidean distance is the metric used for distance calculation in LOGEXP tree; L_k_-norm (k=0.3) for trees in HTML-TM [15]. t-SNE [23] dimension reduction was applied for the 2-dimension visualization of vectors in examples.

### 2.6. Programming

All the TM approach was developed in the AIBIA lab and adapted for this specific study. The supporting tasks, including constructing HTML-TM files, were programmed mainly in Matlab ® [24].

### 2.7. Mapping the literature

The phylogenetic thee with all words in the WRD set is available in the supplementary. Studying this diagram, we performed our initial analysis and got acquainted with terms relations and their global disposition. These previous interactions oriented us for the first exploratory incursion into Yoga literature. We aborded all the review papers (n=306) available to define a prospecting list of LOGEXP. The list was revised to even i) represent most of the literature and ii) be understandable by the reader. Despite directly clustering data, we chose this strategy to derive a more meaningful structure for a multidisciplinary Yoga researcher. We built a vector from each resulting LOGEXP according to the same role used to embed the words, allowing us to exploit them in semantic searches. The phylogenetic tree of the LOGEXP is in the FIGURE 1. For the name of the leaves, we appended to the LOGEXP text the numbers of hits it brought from TextDoc and TextTitles, respectively. The union of all searches cover 99.86% of the documents and 90.58 of the titles in TextDocs.

**Figure 1.**
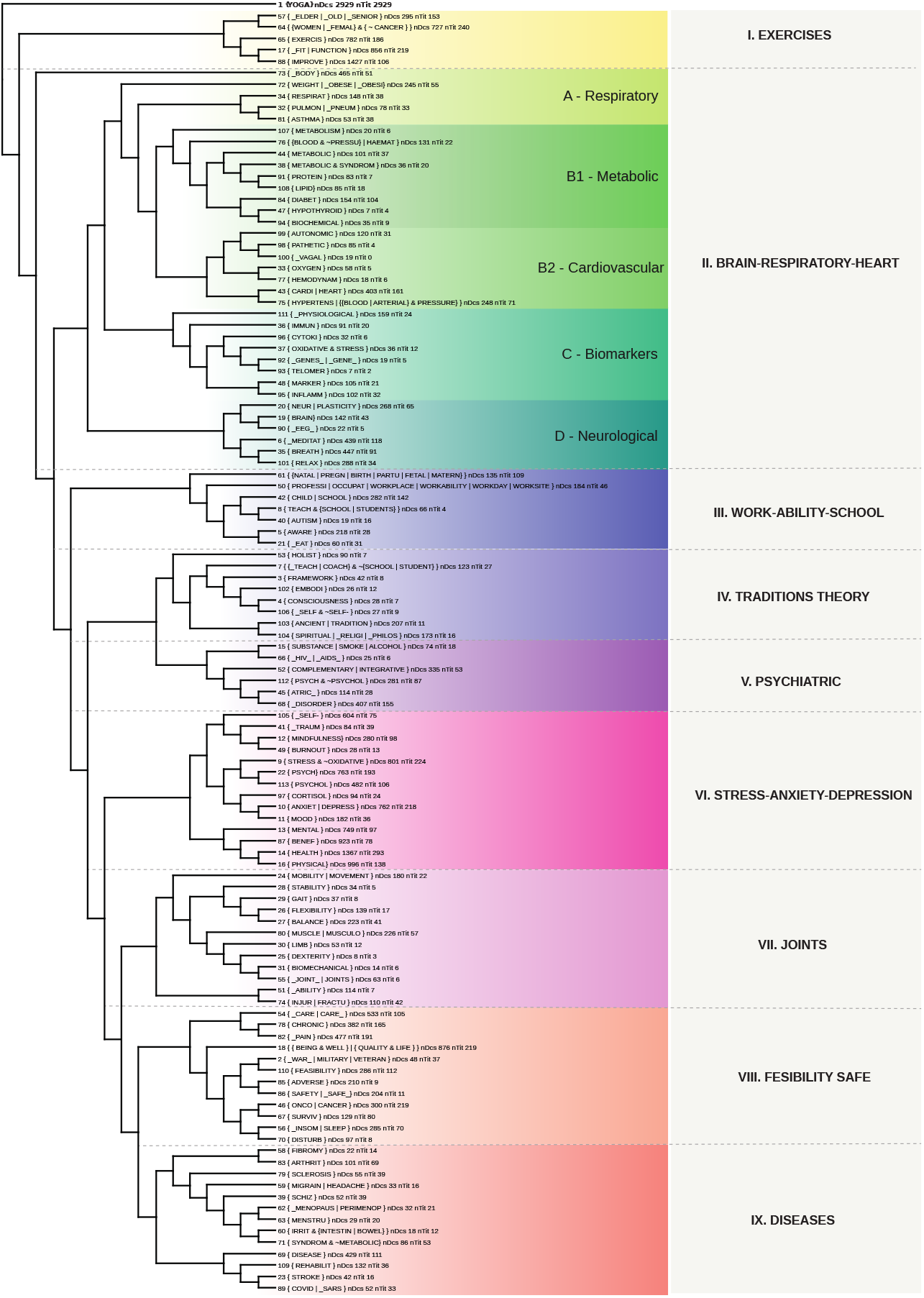
Mapping of Yoga literature with all 113 LOGEXP studied. The phylogenetic tree presents the relationship of the 113 LOGEXP and divides the findings into nine groups with related topics. The tree given characterizes the field of yoga as it has been studied in the literature. Each branch presents each LOGEXP and the associated identification number. It also indicates the number of studies/documents (nDocs) and the number of titles (nTit) related to the logical expression. Group II. BRAIN-RESPIRATORY-HEART is further subdivided into four groups: A. Respiratory, B1. Metabolic, B2.Cardiovascular, C. Biomarkers, D. Neurological.

### 2.8. TM Results

We employed in this study a proprietary TM arsenal aiming freely to adapt resources to our purposes and to tune vector configurations. FIGURE 1 shows an intuitive example of comparing three searches performed by related words in the context of all words (WRD0).

In the topic, literature mapping shows the effectiveness of our strategy and its semantic power. It presents a phylogenetic tree within the LOGEXP rooted in the term ‘YOGA’ (FIGURE 1). We also projected a no-cost way to make these resources available to Yoga researchers in HTML-TM preprocessed files. This environment will even allow the users to access this paper’s results and perform semantic searches in the knowledge base. Phylogenetic trees, vectors, and corresponding generating data are available. A reader more interested in vector representation or word embedding can find vectors in supplementary.

## 3. Literature Mapping

The LOGEXP vectors allowed us to perform semantic searches for mining correspondent relevant literature. In addition, the phylogenetic tree within all prospecting LOGEXP gave us clues to separate them into nine related groups, I-IX (FIGURE 1).

It is important to note that the nine groups are not necessarily mutually exclusive. On the contrary, they are interconnected and can be seen more as characteristics of the literature rather than disjoint groups of articles. However, we seek to explain each of these groups by studying the most relevant semantic search results (papers) in an orderly fashion by the vector at the centroid of each group, complementing them with other forays into the remaining literature when necessary.

### I Exercises

The practice of yoga em alguns casos podem ser considerado is considered a light to moderate impact physical activity; due to its characteristics that aim at a conscious approach, it generates safety to exercise and rehabilitate the body systems. Furthermore, it is considered an exercise that promotes physical, emotional, and neurological benefits, aiming at well-being and quality of life.

The literature has demonstrated the results of yoga practice in health promotion when compared to other physical activities or no physical activity over the years. Older adults, obese people, and individuals recovering from surgeries, traumas, and non-communicable diseases are the main groups that seek YP, aiming at it as a means to adjust physical functions. Regarding gender, the female public predominates in this bias, with women dominating the studies related to the group I.

There are systematic studies that compare YP to conventional physical exercises. In [25], the authors compared yoga practice to aerobic exercise for reducing anxiety symptoms, and the conclusion was in favor of yoga practice; The study [26] compared the intervention of inspected yoga practice versus physical exercises. They performed at home, through videos, for children with Enthesitis-Related Arthritis (ERA). The results showed that except for the parent-reported quality of life, the other parameters were positive for yoga practice. In the study [27], a 6-month experiment of yoga practice in healthy older people was presented, aiming at the effects of yoga practice on cognitive function, fatigue, mood, and quality of life. There was a significant improvement in the yoga group regarding the quality of life and physical measures compared to the army and control groups.

### II Brain-Respiration-Heart

Group II stands out for housing strongly connected subgroups (Blocks) in the literature. With its analysis, we can get a comprehensive view of the relationships between the action of practice (YP) with internal body aspects. Following the hierarchy of the diagram: A-Respiratory; B1-Metabolic and B2-Cardiovascular; C-Biomarkers; D-Neurological. We divide their presentation into these aspects below.

#### D Neurological

Block D in this group is related to articles addressing the neurological and brain aspects of YP. For example, some reports present EEG and neuroimaging results targeting brain aspects related to Yoga practice [28][29], while other papers interpret from a neuroscience point of view the principles that may support the benefits that the YP entails [4][[8].

In the article by Devi et al. (1986), the authors suggest the relationship between Yoga practice and the regulation of sympathetic and parasympathetic activities through the analysis of several biochemical markers in patients with stress. Recently this theme has strongly emerged, and the volume of evidence proves this hypothesis that can be one of the keys to understanding the group with essential reflections in all related literature.

Other works highlight that the improving tone of the parasympathetic system (PNS) and the decrease of the sympathetic nervous system (SNS) activity are beneficial for stress reduction [31] and the benefit of cardiovascular systems functionalities [32]. Those, in turn, are related to several diseases highlighting diabetes (LOGEXP-43, 84) that also appear highlighted in this group. The regulation of ANS is measured through several parameters. Heart Rate Variability (HRV), present in many studies, deserves attention. In addition, higher parasympathetic activity is seen when the High-Frequency component (HRV-HF) predominates. Studies use this characteristic and HRV measures to verify the effect of Yoga practice quality by comparing experienced practitioners with beginners or non-practitioners [33][34][35].

#### A Respiratory

The same techniques used for meditation in traditional YP can be used to promote relaxation and for training attention focus [36]. Breathing techniques for achieving states of meditation or relaxation are widespread throughout the Yoga literature. Self-perception, or awareness (AWARENESS), represents the active posture of the practitioner during exercises and is a link that relates YP to nervous system functioning. Awareness, as understood as ‘being present,’ is also used to learn how to improve the ability to maintain attention [37].

#### B2 Cardiovascular

We kept the search performed by the expressions (LOGEXP-81, 32) in the diagram of FIGURE 1 to maintain it in the context related to breathing (LOGEXP 34). Breath control learned by yoga practitioners also affects the regulation of ANS and the cardiovascular system [38][39]. Papers related to the cardiovascular system frequently address cardiovascular diseases and their risk factors [40][41][42]. In addition, risk factors are usually targeted in studies focusing on hypertension, diet, overweight, and others. Other studies address the regulation of the cardiovascular system by exploring its relationships to the parasympathetic system [38][43][44] (FIGURE 4). The autonomic nervous system appears literally in block II-C, represented by (LOGEXP-98, 99, and 100) (FIGURE 2.B); we realize that understanding the ANS and relationships in group II is fundamental to understanding the Yoga-related literature and is a crucial element of the presented structure. Unfortunately, the LOGEXP-77 (HEMODYNAM) brings only 18 documents. Still, it shows the capacity of the Data Mining Model to infer relations that are not explicit, as it happens in [45]. It suggests the connection of Yoga with hemodynamic adaptations, parasympathetic activities, and block B2 (FIGURE 4).

**Figure 2.**
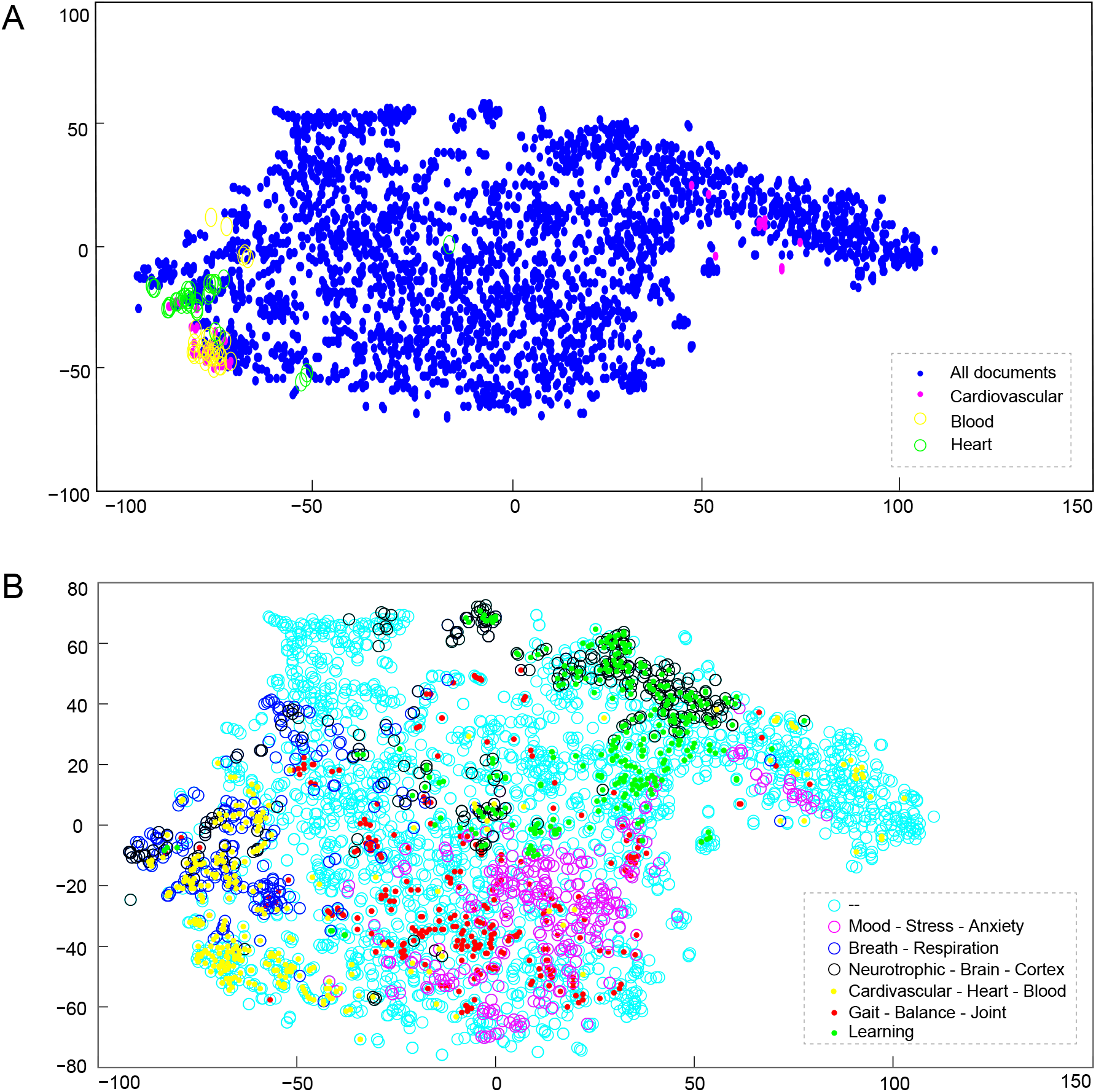
t-SNE visualization of vectorized documents. Each dot represents the TITLE+ABSTRACT for each document. **A**. Semantic searches for HEART (green), CARDIOVASCULAR (magenta), and BLOOD (yellow). The search considers 40 hits closest to each query. **B**. Distribution of 300 nearest hits for six semantics searches listed in the legend. Note that in **B**. the overlap between the topics of ‘NEURO’ (black) and ‘LEARNING’ (green) is visible, as well as between ‘BREATH’ (blue) and ‘CARDIO’ (yellow), and more subtly between ‘MOOD’ and ‘BALANCE’.

#### B1 Metabolic

Block B1 appears strongly related to biochemical markers, and papers showing positive effects of YP [46][47][48] stand out, although the need for further studies is often reported. In this context, we find in the literature connections between Diabetes, Metabolic Syndrome, and even between these and Hypothyroidism. For example, LOGEXP 84 (DIABET) found 154 articles related to YOGA and diabetes, of which 104 ‘DIABET’ appears in the title. Besides pointing out the benefits Yoga can offer directly to practitioners with diabetes, the literature correlated to it, and block II opens perspectives for understanding the physiological aspects of YP.

#### C Biomarkers

The literature presents experiments that relate measurable markers - biochemical, genetic, etc. - to the physiological benefits of YP, which acts particularly in regulating the immune system. In [1], a review of the main results highlights the benefits of YP in various aspects. The authors propose there is a pattern for the regulation of pro-inflammatory markers related to YP. In specific, they highlight the reduction in IL-1beta, IL-6, and TNF-alpha (pro-inflammatory cytokines). In the same study, we found that the favorable effect on hypertension may not be immediate, indicating a lasting impact that may be susceptible to the dosage of the practice. We consider that the effect cited may be related to a learning process consolidated with elaboration in the post-practice period. The authors themselves highlight the possible involvement of the sympathetic nervous system in the downregulation of immune-related parameters described in the literature that may be related to the learning we mentioned. More recent studies confirm this anti-inflammatory feature of YP and add other markers [49][50]. Other studies investigate whether YP can positively influence reducing cellular aging [51][52] and propose telomerase and chromosome length as indicative measures. Studies also address checking markers that show reduced oxidative stress when related to YP [53][54][51].

Other markers positively related to YP are the neuroplasticity marker Brain-Derived Neurotrophic Factor (BDNF) [55][56][57] and to Gamma-Aminobutyric-Acid (GABA), whose low levels are found in severe depression patients. We highlight some among many other markers whose relationship with Yoga practice has been significantly established. (TABLE 1) [3], adapted from [3], brings a list that includes additional features. In this same study, other markers are presented. We noticed, analyzing the results, that many of these markers are also correlated, which suggests a possible pattern to be applied to model generalized YP results evaluation, which, however, demands specific studies.

**Table 1.**
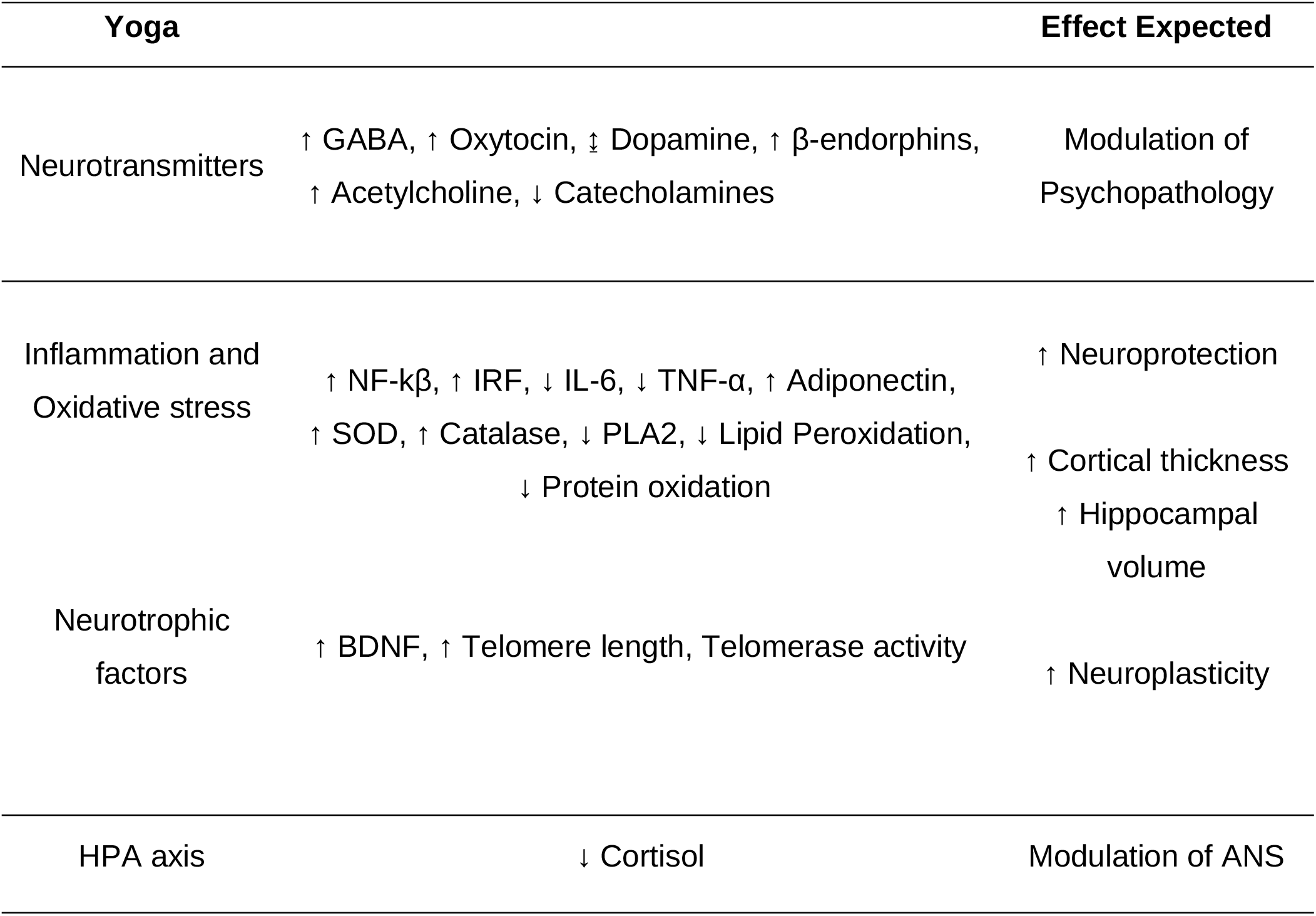
The table presents mapped relations between YP and positive neurobiological effects. The diagram shows important markers integrating with the other studies surveyed. Adapted from [3].

### III Work-Ability-School

The studies mostly related to this group address specific social contexts, such as school and work environments, where the insertion of yoga (and Mindfulness)-based programs are applied and tested. The results, in most cases, are obtained through qualitative tests. The studies often involve groups of students at various levels [72][73][74][75] and teachers [76][77]. The studies target situations where the YP can generate benefits in several aspects, such: Stress, anxiety, depression, resilience, self-esteem, compassion, social relationships, academic performance, mindfulness, flexibility, communication, empowerment, physical fitness, and self-regulation skills in various forms (emotional, relaxation, mindfulness, aggression, etc.). As an example, YP helps regulate emotions in students [75][78] and improves the ability of children on the autism spectrum to communicate their feelings [79][80]. Other settings, such as pregnancy groups, healthcare settings, and vulnerable communities, also host studies with similar characteristics. In addition to direct benefits to practitioners, feasibility and adherence to programs are also evaluated [81][82][83]. An exciting example of the application of Yoga in the academic environment is its use in teaching anatomy, where students use self-perception as a guide for learning [84][85][86]. Another notable aspect is the maintenance of effects after Yoga programs are ended [87][81]. These findings reinforce the learning effect related to the benefits of yoga practice.

### IV Traditions-theory

Group IV represents articles that establish theoretical connections between Yoga and science or philosophy. In them are approached concepts of traditional knowledge in Yoga practice establishing relationships with contemporary science.

Concepts of self-image and embodiment stand out in theoric studies about Yoga in Psychology. Some examples are papers concerning obesity [10][88][7][89]. However, our analysis found elements in Embodied cognition theory that can corroborate the elaboration of new frameworks and strategies to design studies about Yoga.

The analysis of social contexts in which practitioners may be included is addressed in [90] and [89]; the relevance of the benefits that self-care based on yoga techniques offers to accessibility are discussed in [91]. Finally, the use of YOGA in work settings and interpersonal relationships is contextualized in [92] and [93].

Ethical aspects are present in works that approach traditional principles of Yoga in articles in this group, like [94].

The Theoretical approaches in neuroscience are likely the best representative for this group. The recent emergence of studies aims to explain or propose ways to uncover why Yoga practice produces the benefits observed. [8] relates YP and POLYVAGAL theory. In this work [8] and [4], we notice the relevance of the association between Yoga practice and the regulation of the autonomous nervous system, which occurs mainly by improving the tone of the PNS. The scientific studies that seek to substantiate the principles of Yoga practice are of great relevance in the current scenario where the literature is fragmented in different contexts and diverse approaches and are essential to better understanding and exploring Yoga techniques.

However, we still notice that new contributions with a multidisciplinary scope that can integrate Yoga knowledge in several areas of expertise are essential. [4] proposes three components to be considered in describing Yoga practice - POSES-ATTENTION-BREATHING; these are three active forms of involvement that, as indicated by literature, bring benefits at Yoga work. We observe, however, that these elements, when acting together, can result in lasting benefits that we associate with another possible component: ‘LEARNING.’ We believe that the systematization of new structures for Yoga should explicitly contemplate Learning as one of its fundamental principles.

### V Psychiatric

With many indications of benefits, Yoga is a promising source of support for treatments that can add positive effects to medicine. However, the complexity of the area and the methodological limitations of the studies make it difficult to make safe prescriptions in medical procedures and practices [95]. Many studies fail to be conclusive despite noting often positive results. We observed that the literature is more assertive when articles deal specifically with Yoga concerning those that approach the theme in a joint context with other Complementary and Alternative Medicines CAM comes. Therefore, in the definition of the LOGEXP we considered only articles that contain the term YOGA explicitly in the title. Furthermore, since we aim to understand Yoga literature and not necessarily to substantiate Yoga-based therapies, we intend to understand the motivations of the studies and the relationships between them to point out new perspectives that contribute to the field.

Group V approaches Yoga as a complementary therapy aiming at psychiatric disorders, including eating and addictive disorders. The article [3] highlights the evolution of Yoga research in psychiatry that, from works that aimed only at the general well-being of practitioners, began to be approached to deal with better-formalized aspects, taking into account neurobiological effects, measurable and relevant to medical practice. It also highlighted relationships pointing to important markers with mention of the direction of the observed effect and its relationship to YP - positive aspects expected in possible interventions. The diagram shown in TABLE 1 [3] is a summary that can also be used to integrate this topic with others in this study, so we consider it essential to adapt and integrate it into this study. Furthermore, many markers in the diagram have already been presented in the discussion of group II, reinforcing the possibility advanced, that YP-related characteristics also correlate analogously with each other. If we abide by this principle, we can trace the possible relationships by traversing the evidence provided by the markers on the various research fronts, inferring relationships helpful in understanding the area. These efforts are beyond this paper’s scope, but we think it is a promising avenue for further studies.

### VI Stress-anxiety-depression

While we found in group IV elements that highlight the psychiatric aspects of YP, in group VI, the emphasis is on the psychology field where affective, mood, social relations, overload, and trauma issues are treated. Some works place Yoga as a potential adjuvant in therapeutic processes or even as a possible substitute in some cases with advantages related to accessibility [96][97][98]. In the yoga literature, LOGEXP-113 (PSYCHOL) is mainly associated with stress in addition to anxiety and depression. LOGEXP-10 (ANXIET | DEPRESS) finds more than 25% of the documents and appears in 218 titles. These numbers show that group-related aspects are present in almost all the literature.

MINDFULNESS is strongly related to the articles representing the group and has different connotations. Sometimes it is taken as the state of the present mind (or the ability to achieve it) [99] and sometimes as a technique to be applied in association with Yoga (MINDFULNESS) [100]. The state of MINDFULNESS appears associated with the practice of Yoga and its positive effects, lower stress-anxiety-depression rates, for example.

Other studies show the benefits of YP in groups with post-traumatic stress disorder (PTSD): It improves mindfulness among other uses in veterans [81][101]; acts in symptoms in women with PTSD [102][103]; and others.

Several studies associate the decrease in cortisol levels related to YP and the reduction of stress and other mood-related disorders. This finding makes us consider the possibility of including mood-related issues and other psychological aspects in the relationships with other markers that have already been addressed, opening new research fronts.

### VII Joints

The sub-items in group VII are related to the studies involving the benefits of Yoga in the neuromuscular system. Unfortunately, we have not found any study that contains a systematic view integrating the aspects of group VII in the yoga literature. However, we present some examples of studies with the features of the group.

The study [104] measured the range of motion of ankle joints in yoga practice, aiming to have a parameter to contribute to the discussion about the expected effects of YP after injury or surgery. The case study [105] applied Yoga to an individual with an incomplete spinal cord injury. The result was an improvement in the following aspects: balance, flexibility, muscle strength of hip extensors, hip abductors, and knee extensors, and improvement in functional goal performance. Another study[106] evaluated the effect of a yoga program to minimize knee adduction moments for women with knee osteoarthritis, and the results showed improvements in pain reduction, strength, and mobility. Finally, a recent [107] described the effect of yogasanas intervention on balance performance in people with Peripheral Diabetic Neuropathy - (NPD). Comparing yoga intervention with conventional exercises, the first was more effective in improving static and dynamic balance performance in all variables of standing balance and reduced fear of falling among individuals with NPD. Other related studies address: physical function [108][109]; musculoskeletal rehabilitation; mobility [110][111][112]; balance [111][113]; gait [111]; flexibility [112].

The hypothesis that yoga generates learning also applies to improving the effects of strength application and biomechanical organization of the body as a whole. The individuals with healthy joints deliver their body mobility, stability, flexibility, balance, dexterity, and gait quality and can improve the effects of biomechanical strength. The organized body system helps maintain focus, enhances coordination, provides neurological reconnections (FIGURE 3.B), and regulates muscle tone and strength, among other aspects. This health condition certainly implies benefits in regulating other systems as well. However, we did not find studies that focus on joints systemically as a criterion for assessing YP. We noticed room in the literature for contributions along these lines.

**Figure 3.**
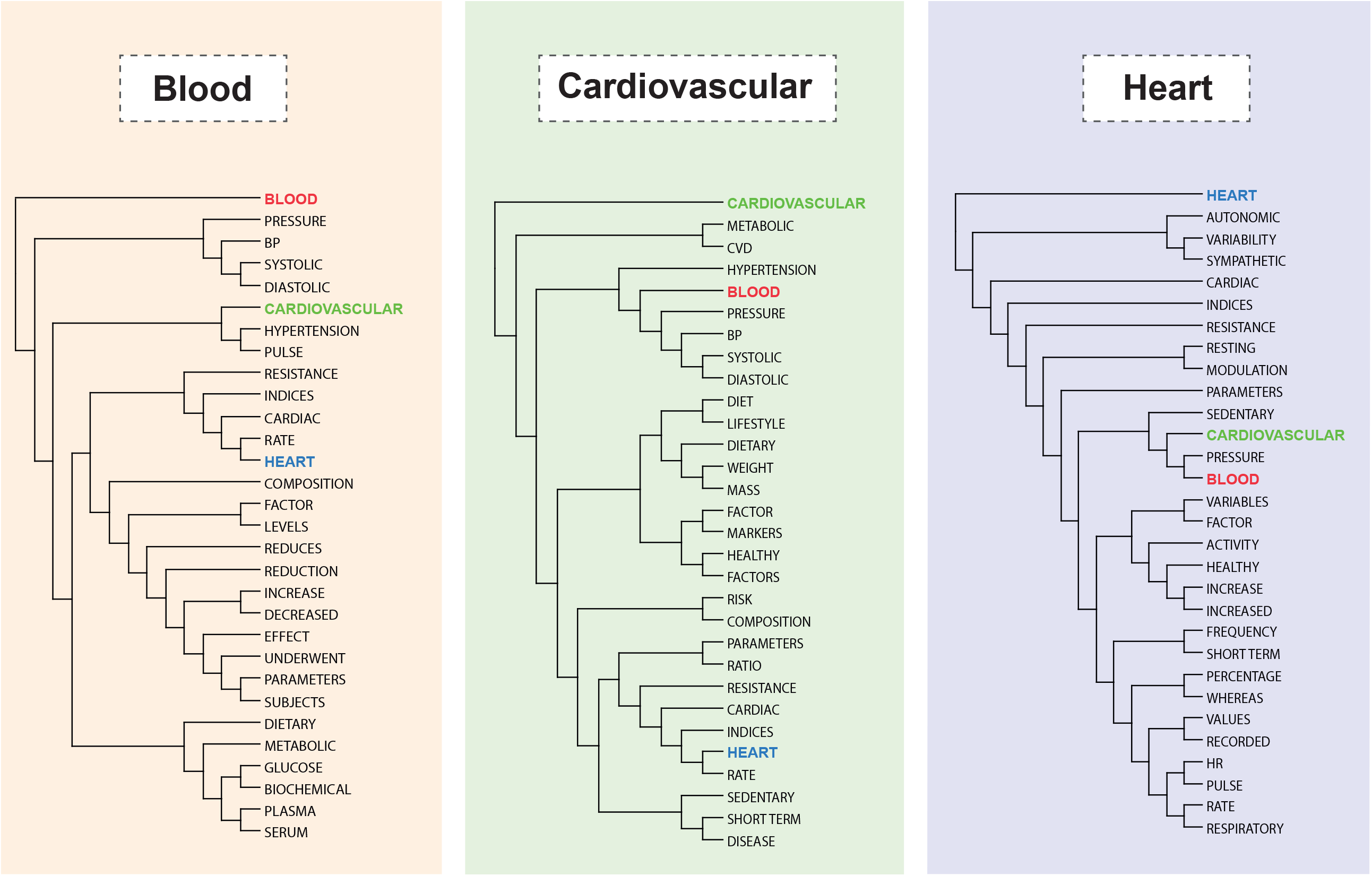
Relations of top results in semantic searches for the words BLOOD, CARDIOVASCULAR, and HEART in the set of all words. These representations demonstrate the semantic capacity of the method to relate the terms. The trees are consistent with the t-SNE in FIGURE 2.A, indicating the relationship among the concepts (close or overlapping points in the t-SNE 2). For example, in both t-SNE and trees, ‘BLOOD’ and CARDIOVASCULAR’ are most relative to each other and more distant from ‘HEART’.

### VIII Feasibility-Safe

People who require special care are the target of programs that search for practices that allow them to obtain a better quality of life and better social insertion. However, It is not always possible to use mass resources to serve the public. In the literature, and particularly in this block, there are studies analyzing the viability of integrating people with particular needs related to diseases in Yoga groups and the possible personal and social benefits that this can bring.

There are frequent works that aim at groups of recovered cancer patients [114] [115] or in treatment [116][117][118]. Other papers address groups of people suffering from chronic pain that demand therapeutic practices adapted explicitly to them [119] [120]. The expected outcomes in the programs proposed in these studies involve improving the quality of sleep, mood, anxiety, and depression; relieving stress; reducing fatigue; increasing participants’ physical and social functionality; decreasing suffering overall in addition to others. However, the studies of this group are not only limited to seeking the well-being of the participants but also show that Yoga-based therapies can play a relevant role in filling the gap that conventional methods cannot address. The program’s viability is checked with criteria such as effectiveness, recruitment, adherence, participation, safety, and satisfaction.

One of the features of the YP is the conscious and focused movement, which provides active and personal participation of those involved in the activities. Therefore, the care in performing the practice promotes a reduction of risks and adverse effects, which we believe may motivate a part of these studies.

### IX Diseases

Although group IX presents a heterogeneity of diseases and syndromes, the fact that the group has been organized cohesively makes us assume a tendency of common elements in the literature that make them aggregate in this block. The articles describe a certain peculiarity regarding YP with positive results concerning relieving pain or suffering caused by the listed diseases. We comment on some of the related works below.

The literature shows that yoga interventions can be a valuable way to rehabilitate individuals with acquired brain injuries such as stroke and neurodegenerative diseases such as Parkinson’s and Multiple Sclerosis and are included in this promising role of benefits to cardiovascular rehabilitation [121] [122] [123] [124]. The main results reported are: a) improvement in cognition, mood, and stress reduction; b) increase of self-efficacy and motivation for physical activity; c) Motivation for physical activity; d) improvement in quality of life; e) better subjective perception of cardiac health, with a tendency to improve left ventricular systolic function; and f) improvement in motor function in balance. Concerning balance, a study [113] mentions that individuals who practice yoga longer show better balance control. This result reinforces that yoga practice creates learning memories.

The articles show that yoga plays a crucial role in system rehabilitation. The studies [125] and [126] corroborate the benefits of yoga practice in relieving osteoarthritis-related pain.

Regarding LOGEXP-59 (Migrain | Headache), the study [127] showed that yoga practice resulted in clinical improvements, reducing headaches’ frequency and medication intake. It also observed an increase in the vagal tonus with a reduction in sympathetic activity, improving the cardiac autonomic balance.

Because fibromyalgia is directly related to various types of pain and has a direct relationship with joint pain, the link in the pain role of this group is justified. One study [128] reports that yoga and mindfulness practice improve fibromyalgia manifestations, functional impairments, and coping skills, modulate abnormal fibromyalgia-related pain processing, and significantly increase heat pain tolerance and pressure pain threshold.

The study in [129] concluded that yoga and meditation significantly improved symptoms of stiffness (plus anxiety and depression), improving well-being.

Once again, pain is present in the literature as a link between the LOGEXPs of group IX when we analyze LOGEXP-60, which refers to Irritable Bowel Syndrome (IBS).

The study [130] describes the main symptoms of (IBS): abdominal discomfort, diarrhea, and constipation; emotional stress seems to exacerbate the signs of the syndrome, being a common denominator among the participants of the study conducted. The study [131] - Systematic review suggests that yoga may be a safe adjunctive treatment for IBS. Other literature [132] - Narrative review of technologies - Evidenced the effectiveness of yoga as a therapy for IBS.

One of the symptoms that can affect women in their reproductive period is primary dysmenorrhea, which is cramps in regular menstrual cycles, not associated with more severe problems. The study [133] evaluated the effect of yoga on aspects related to pain relief in the menstrual period, improving physical fitness, and quality of life. The quality of life and balance of post-menopausal osteoporosis was also evaluated in [134]. The study concluded that Yoga is an alternative for pain management with applications in physical and social rehabilitation in individuals with osteoporosis. In addition, the study [135] concluded that Yoga positively contributes to women’s physical and psychological quality of life in the perimenopausal period.

Yoga is a practice of a safe nature. Their characteristics stimulate the adhesion to Yoga groups aiming for physical activity. In addition, people with physical, emotional, or psychological challenges resulting from diseases or central nervous system disorders are an essential part of this public. Literature shows positive aspects of mind-body yoga, meditation, and relaxation techniques for individuals with chronic and debilitating neurological diseases such as Multiple Sclerosis, Stroke, or Parkinson’s. In studies related to Yoga practice and central nervous system disorders, it is reported that yoga practices can be safe and effective for quality of life, balance ability, gait pattern, and joint flexion [5][136]. However, other articles claim that due to the heterogeneity of yoga practices, and the lack of protocols or outcome indicators for practices targeting specific audiences, there is insufficient evidence confirming the benefit of yoga compared to other physical exercises.

People have a natural capacity for adaptation that includes reconnecting the poorly explored and injured areas of the body. We conjecture that Yoga techniques aim to explore these mechanisms efficiently, enabling physical, emotional, and neurological benefits. Unfortunately, exercise-based rehabilitation programs are usually expensive, while yoga has presented a safe and low-cost alternative.

## 4. Discussion

The search for the PARASYMPATHIC Figure 4, for example, associates elements of the nervous, cardiac and respiratory systems, highlighting the relationship that the literature establishes between them and Yoga, the search area. On the other hand, the search for related articles, performed using the vector of a keyword, brings us closer to pieces that are important to delve into related topics. Moreover, semantic investigations reveal aspects that other approaches may not allow.

We can infer that an article is related to the search term even when it is not explicitly present in the text. For example, this is the case of [58], the first result of the search for articles using PARASYMPATHIC; however, the search term is not present in the corresponding document (Figure 4).

**Figure 4.**
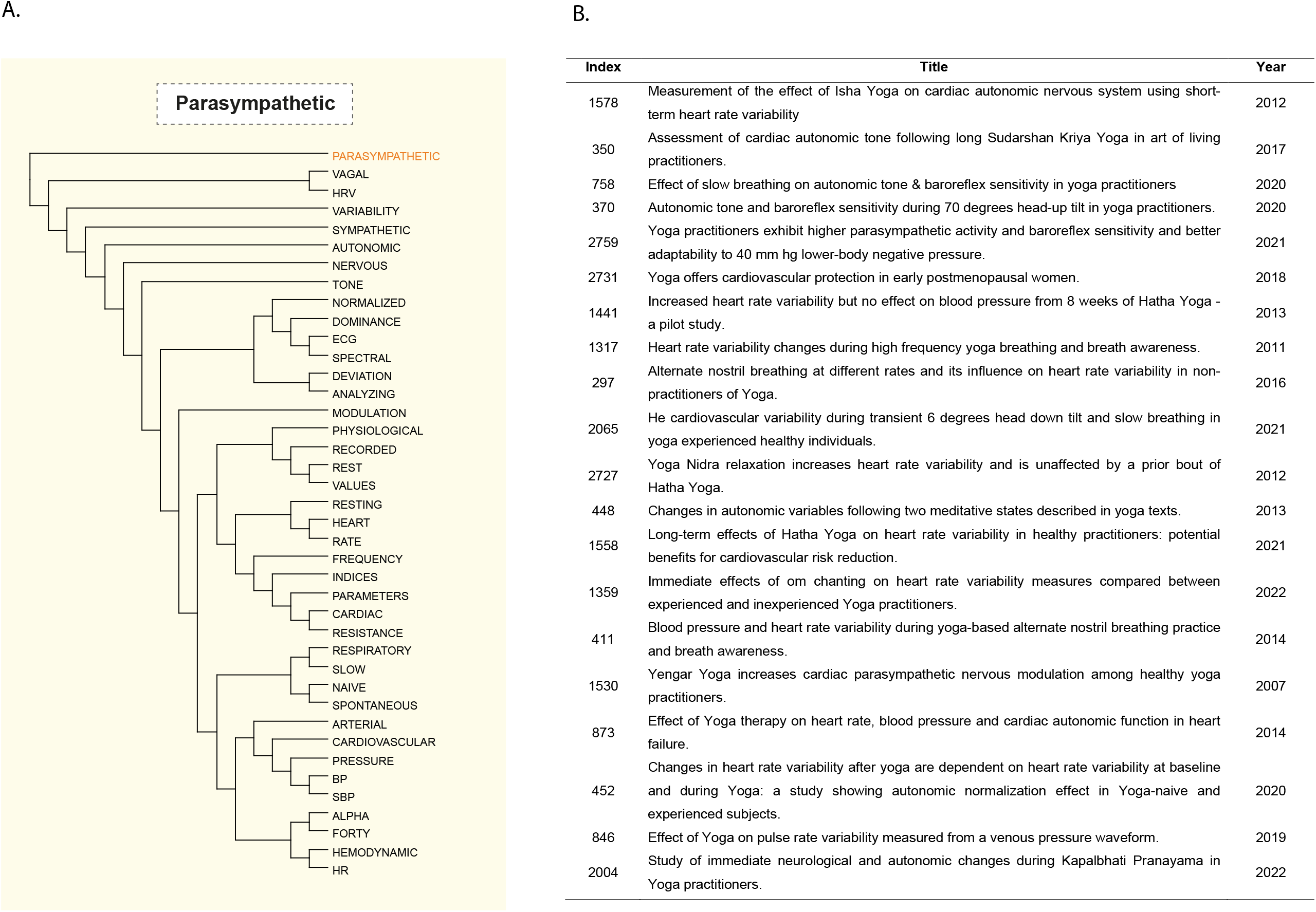
It is possible to obtain the closest words or studies within the same vector space from a vectorized word. From the word ‘PARASYMPATHETIC’ as an example, we can receive: **A**. phylogenetic tree associating the word with the 40 closest words according to Yoga literature and **B**. list of the closest papers to the word. Note that the tree shows words related to the subject, such as ‘NERVOUS’, ‘AUTONOMIC’, and physiological characteristics modulated by the PARASYMPATHETIC system, such as ‘HEART’, ‘RATE’, ‘CARDIAC’, ‘PRESSURE’, ‘HEMODYNAMIC’, related to the cardiovascular system, and ‘RESPIRATORY’. Similarly, the listed papers present the cardio-respiratory and autonomic nervous systems as the central subject of the study. The references for articles are [58][59][38][60][45][44][61][62][63][64][65][66][67][35][68] [69][43][33] [70][71]

To build the mapping, we designed 113 textual expressions for searches in the literature (LOGEXP) and coded them in vector representations to enable semantic searches. The LOGEXP were then automatically organized hierarchically Figure 1. Based on the associations obtained, we could discuss the literature by dividing it into nine characteristic topics and maintaining a line of integration between them. With these nine cohesive groups we are able to outline and understand the major aspects of the yoga field of study. We always seek to validate the observed relationships between words that are affected by the context of the literature.

The relationships were coherent, and we showed diagrams where searching for a word reveals its context, making it easy to delve deeper into the literature. Although multifaceted, the literature is consistent, and we can trace the predominant elements even among its more distant aspects.

We visualized the possibility of organizing the markers - positively related to YP - to eventually describe the health state associated with it; However, this elaboration is beyond the scope of this paper, and though it is an instigating possibility. Another aspect that caught our attention is the weak representation in the analyzed literature of terms that we consider essential in the context of Yoga. Therefore, we will comment on only two of them.

JOINTS (also associated with JOINT) appear almost only in situations related to trauma or pathologies. We did not find any study that approaches joints in a systemic way that evaluates, for example, preventive benefits or the gain promoted by the health of the joints with mobility, flexibility, and the other aspects mentioned in group VII. Yoga’s quality gain in joints may improve the known correlations to other elements found in the literature and could be better explored in future studies.

LEARNING was eliminated from the list of relevant words and is not among the words in HTML-TM (WORDS); this is because the frequency of the term in Yoga literature is lower than that observed among words in general usage. We included two searches involving the concept of ‘Teaching’ in the set of logical expressions (LOGEXP-7 and LOGEXP-8). While LOGEXP-8 targets relationships in the school environment, LOGEXP-7 targets are teaching outside the school context ∼(SCHOOL | STUDENT) and, therefore, the teaching of Yoga itself. In this second context, we realize its position is given within group IV-Traditions-Theory. Traditionally there are Yoga schools, and the presence of experienced instructors is the norm in YP. Much, if any, attention is given to teaching the practice in the literature. Although some works emphasize the learning effect resulting from YP in preserving the results obtained, the gain of resilience and physical, mental, emotional, cognitive, and other skills [137][138] [139][140][141]. The semantic relation of LEARNING is also highlighted in Figure 2-B, where we can see the concept is closely related to Neural-Brain aspects.

However, the literature does not include learning aspects as a structural element for studying Yoga. We believe there is a need for studies that explicitly engage learning as one foundation of Yoga in the contemporary context. One aspect that can be considered, for example, is the study of learning curves related to the parameters identified as positive results of YP.

Finally, we believe that the available material helps Yoga researchers promote the multidisciplinary interconnection of investigations with the consequent contribution to the area’s evolution.

## Data Availability

All data produced are available online at:
https://drive.google.com/drive/folders/1OYgY33lirN3fuo5HttvWLHWc2EH13Mwu?usp=sharing

## Data availability

The data obtained from the analyses performed in this study are available at: https://drive.google.com/drive/folders/1OYgY33lirN3fuo5HttvWLHWc2EH13Mwu?usp=sharing

## Acknowledgments

The authors thank the group of Artificial Intelligence Applied to Bioinformatics of Federal University of Paraná.

## Author Contribution

R.I.R. and R.T.R. designed and implemented the analysis. D.J.M. contributed to the bibliographic survey. R.I.R. and R.T.R. wrote the original draft of the manuscript. C.R.D.P, C.P.P, and J.N.M. made substantial contributions, revisions and approved the final manuscript. R.T.R. supervised the whole project. All authors contributed thoughts and advice, discussed the results, and contributed to writing the final manuscript.

